# COVID-19 booster vaccine attitudes and behaviors among university students and staff: the USC Trojan Pandemic Research Initiative

**DOI:** 10.1101/2021.12.09.21267545

**Authors:** Ryan C. Lee, Howard Hu, Eric S. Kawaguchi, Andre. E Kim, Daniel W. Soto, Kush Shanker, Jeffrey D. Klausner, Sarah Van Orman, Jennifer B. Unger

## Abstract

**Introduction:** Although COVID-19 vaccines significantly reduce morbidity and mortality, recent evidence suggests that immunity wanes after 6-9 months, and that a third vaccine dose could further reduce COVID-19 transmission and severe illness. However, previous studies have not assessed attitudes about getting booster doses. This study examined COVID-19 booster vaccine attitudes and behaviors among university students and staff in the fall of 2021.

**Methods:** Participants responded to an email invitation and completed electronic surveys. Results. In our sample, 96.2% of respondents indicated willingness to get a COVID-19 booster shot at least once per year. In both bivariate and multivariate analyses higher trust in science was associated with having higher odds of booster willingness. Those who identify as Black, on average, reported trusting science less than other racial/ethnic groups.

**Conclusions:** Our findings demonstrate high willingness to receive a COVID-19 booster shot and highlight the importance of educational and motivational messages that focus on trust in science to increase willingness to get the COVID-19 booster. More research is needed to better understand the impact of cultural beliefs on booster willingness and vaccine hesitancy. This understanding will help determine what messages and populations to target to increase booster willingness in the future.

## Introduction

Although the Pfizer and Moderna COVID-19 vaccines significantly reduce morbidity and mortality, recent evidence suggests that immunity wanes after 6-9 months, and that a third vaccine dose could further reduce COVID-19 transmission and severe illness.^1,2,3^ Numerous articles have reported hesitancy to get the first or second dose.^4,5^ However, previous studies have not assessed attitudes about getting booster doses. Hesitancy to get booster doses could result from reluctance to get vaccines frequently, severe side effects from previous vaccines, and concerns about the fairness of getting a third dose when other people lack access to any doses.

As COVID-19 boosters become more widely available to the general public, information about attitudes toward third vaccine doses could inform the design of health communication messages for more hesitant groups. Using data from an ongoing survey of university students and staff, we assessed attitudes toward third vaccine doses and the demographic correlates of these attitudes. We hypothesized an overall high willingness to get a COVID-19 booster shot because the sample was from a population of college students and staff/faculty in Los Angeles, where vaccination rates were relatively high. However, we also hypothesized some sub-groups may have lower booster willingness compared to others. For example, the high level of vaccine hesitancy among Black Americans (referred to as Blacks in the remaining part of this paper) may suggest there will be a lower booster willingness as well.^6^

## Methods

### Participants

The participants in our study were students, staff, and faculty at the University of Southern California (USC) in Los Angeles, California. Participants were eligible if they were currently employed or enrolled at USC, were at least 18 years of age, and provided informed consent.

### Procedure

This study was approved by the USC Institutional Review Board. In August - November 2021, we conducted a survey of students, staff, and faculty who were a part of the Trojan Pandemic Response Initiative Health Cohort. We emailed 5,256 students, staff, and faculty (2,876 students, 2,380 staff/faculty) to complete an online survey and received 3,923 responses (75% response rate).

### Measures

The survey assessed demographic characteristics including self-identified race and ethnicity, gender identity, age, and student/staff status. We also assessed self-reported prior COVID-19 infection status and self-reported vaccination status. We assessed respondents’ trust in science using Nadelson’s Trust in Science Scale.^7^ As the main outcome variable, the survey asked how often participants would be willing to get a COVID-19 booster vaccination. Booster willingness was coded as “willing” (any response other than “never”) and “unwilling” (“never”).

### Data analysis

Bivariate logistic regression models were run to examine the marginal effects of demographic and psychosocial correlates on booster willingness among those who were fully vaccinated and completed all the measures of interest (N = 3,668). A multivariable model was run to estimate effects adjusting for confounding by the other variables in the model. We reported unadjusted and adjusted odds ratios and their respective 95% confidence intervals.

## Results

Of the 3,668 respondents, 96.2% indicated willingness to get a COVID-19 booster shot at least once per year, and nearly two-thirds were willing to get boosters as often as necessary (after removing those with missing data) **(Table 1)**. In bivariate analyses, those without prior COVID-19 infections had higher odds of booster willingness compared to those with self-reported prior COVID-19 infection (OR = 1.99; 95% CI = 1.24-3.07), and higher trust in science was associated with having higher odds of booster willingness (OR = 8.36; 95% CI = 6.10-11.60) **(Table 2)**.

**Table 1.** Booster Willingness Among Students, Staff, and Faculty at a Los Angeles University. Responses from 3,668 participants to the question: “How often would you be willing to get a COVID-19 booster, if offered?”. Self-identified gender excludes any “other” category.

|  | **Willing**<br>**(n = 3530)** | **Unwilling**<br>**(n = 138)** | **P-value** |
| --- | --- | --- | --- |
| **Booster Willingness** |  |  |  |
| *As many times as needed* | 2346 (64.0%) |  |  |
| *Three times a year* | 23 (0.6%) |  |  |
| *Twice per year* | 207 (5.6%) |  |  |
| *Once per year* | 954 (26.0%) |  |  |
| *Never* |  | 138 (3.8%) |  |
| **Gender** |  |  | 0.09 |
| *Ref = Female* | 2360 (96.6%) | 82 (3.4%) |  |
| *Male* | 1170 (95.4%) | 56 (4.6%) |  |
| **Age Group** |  |  | 0.92 |
| *Ref = 18-23* | 1131 (96.5%) | 41 (3.5%) |  |
| *24-39* | 1545 (96.1%) | 62 (3.9%) |  |
| *40-49* | 386 (95.8%) | 17 (4.2%) |  |
| *50+* | 468 (96.3%) | 18 (3.7%) |  |
| **Race/Ethnicity** |  |  | 0.004 |
| *Ref = White* | 1220 (96.4%) | 45 (3.6%) |  |
| *Asian* | 1123 (97.7%) | 27 (2.3%) |  |
| *Black* | 186 (93.9%) | 12 (6.1%) |  |
| *Latinx* | 740 (95.1%) | 38 (4.9%) |  |
| *Other/Multiracial* | 261 (94.2%) | 16 (5.8%) |  |
| **Prior COVID Infection** |  |  | 0.002 |
| *Ref = Yes* | 373 (93.7%) | 25 (6.3%) |  |
| *No* | 2933 (96.7%) | 99 (3.3%) |  |
| *Don't Know* | 224 (94.1%) | 14 (5.9%) |  |
| **Division** |  |  | 0.32 |
| *Ref = Staff* | 1550 (95.9%) | 67 (4.1%) |  |
| *Student* | 1980 (96.5%) | 71 (3.5%) |  |
| **Trust in Science Score - mean (sd)** | 4.08 (0.52) | 3.39 (0.62) | < 0.001 |

**Table 2.**
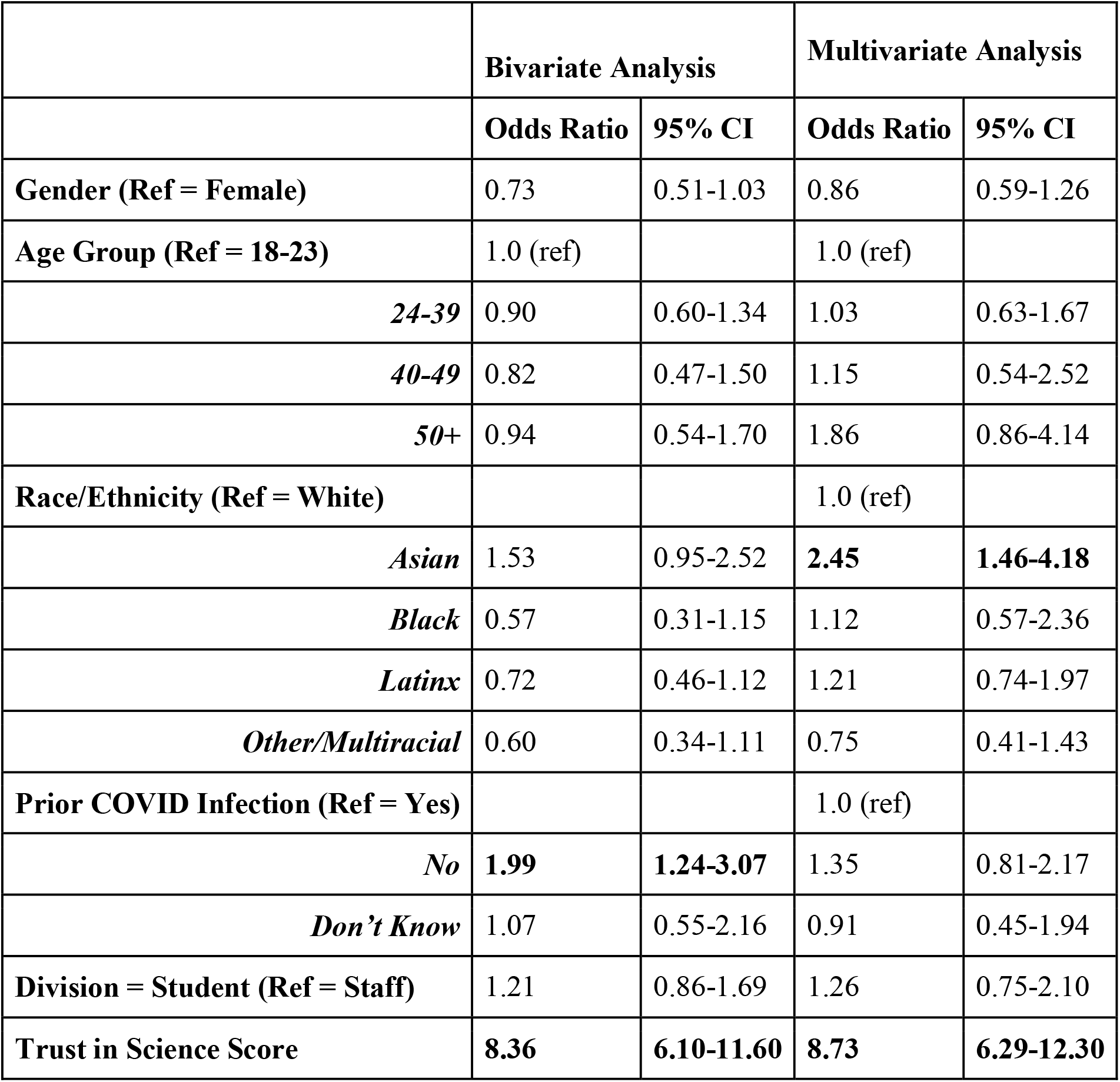
Univariate and Multivariate Correlates of Booster Willingness.

|  | **Bivariate Analysis** |  | **Multivariate Analysis** |  |
| --- | --- | --- | --- | --- |
|  | **Odds Ratio** | **95% CI** | **Odds Ratio** | **95% CI** |
| **Gender (Ref = Female)** | 0.73 | 0.51-1.03 | 0.86 | 0.59-1.26 |
| **Age Group (Ref = 18-23)** | 1.0 (ref) |  | 1.0 (ref) |  |
| *24-39* | 0.90 | 0.60-1.34 | 1.03 | 0.63-1.67 |
| *40-49* | 0.82 | 0.47-1.50 | 1.15 | 0.54-2.52 |
| *50+* | 0.94 | 0.54-1.70 | 1.86 | 0.86-4.14 |
| **Race/Ethnicity (Ref = White)** |  |  | 1.0 (ref) |  |
| *Asian* | 1.53 | 0.95-2.52 | **2.45** | **1.46-4.18** |
| *Black* | 0.57 | 0.31-1.15 | 1.12 | 0.57-2.36 |
| *Latinx* | 0.72 | 0.46-1.12 | 1.21 | 0.74-1.97 |
| *Other/Multiracial* | 0.60 | 0.34-1.11 | 0.75 | 0.41-1.43 |
| **Prior COVID Infection (Ref = Yes)** |  |  | 1.0 (ref) |  |
| *No* | **1.99** | **1.24-3.07** | 1.35 | 0.81-2.17 |
| *Don't Know* | 1.07 | 0.55-2.16 | 0.91 | 0.45-1.94 |
| **Division = Student (Ref = Staff)** | 1.21 | 0.86-1.69 | 1.26 | 0.75-2.10 |
| **Trust in Science Score** | **8.36** | **6.10-11.60** | **8.73** | **6.29-12.30** |

In multivariate analyses, controlling for confounding by the other variables in the model, Asians had over twice the odds of reporting being willing to get at least one COVID-19 booster compared to Whites (OR =2.45; 95% CI = 1.46-4.18) and stronger trust in science was associated with higher odds (OR = 8.73; 95% CI = 6.29-12.30) **(Table 2)**. Notably, we did not detect a significant association between Black race and booster willingness in both the bivariate and multivariate analyses, despite our original hypothesis. To examine any racial and ethnic differences in trust in science, we conducted a post hoc Kruskal Wallace test (p<0.001). Blacks reported lower trust in science than other racial/ethnic groups **(Figure 1)**. Additionally, those who were willing to get a booster trusted science more, on average, than those who were not willing among all race/ethnicity groups **(Figure 2)**.

**Figure 1.**
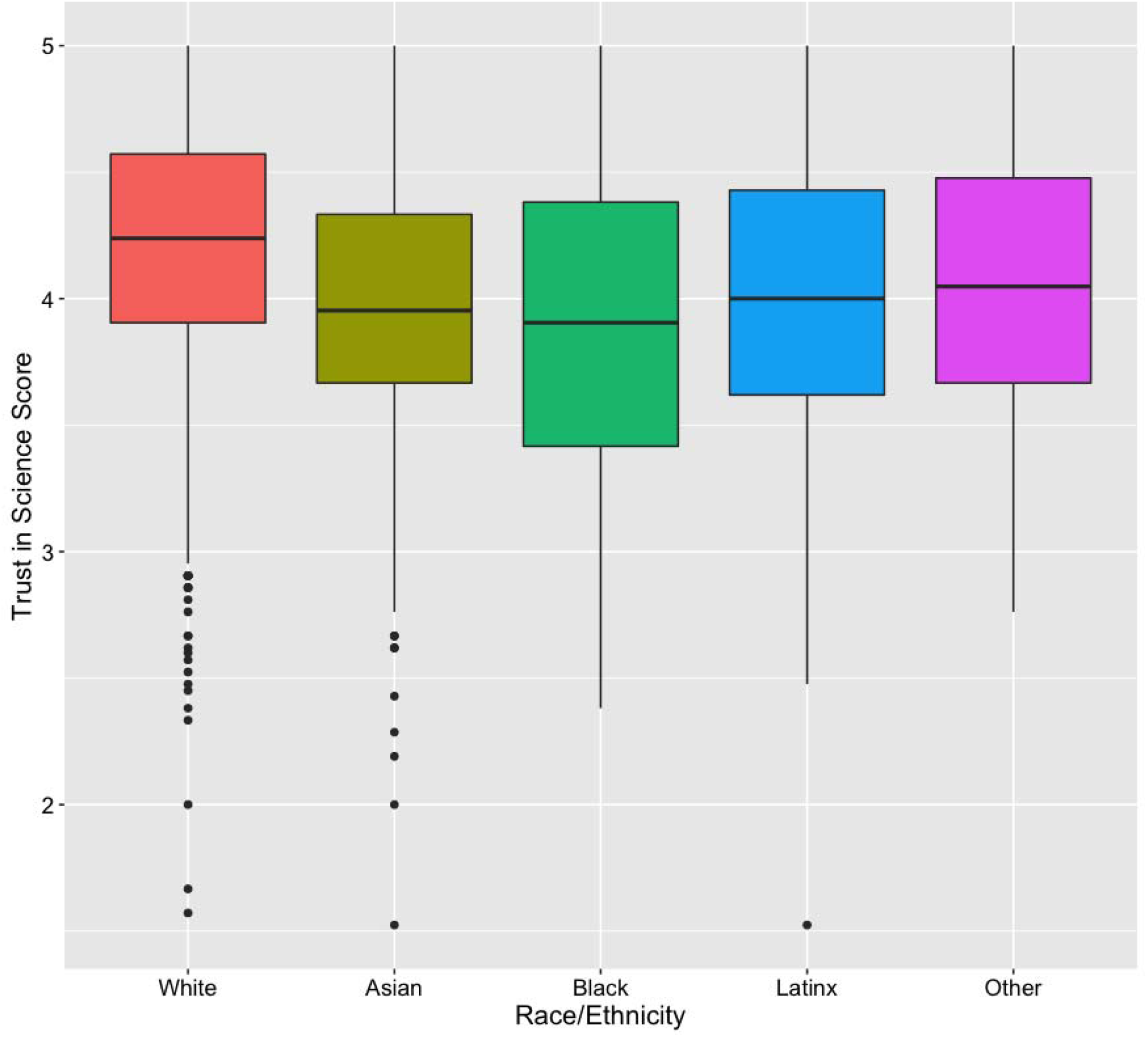
Trust in Science Scores by Racial/Ethnic Groups. Black respondents had a significantly lower mean trust in science score compared to all other racial/ethnic groups (Kruskal Wallis test p<0.001; Blacks vs. All Other Racial/Ethnic groups p<0.001).

**Figure 2.**
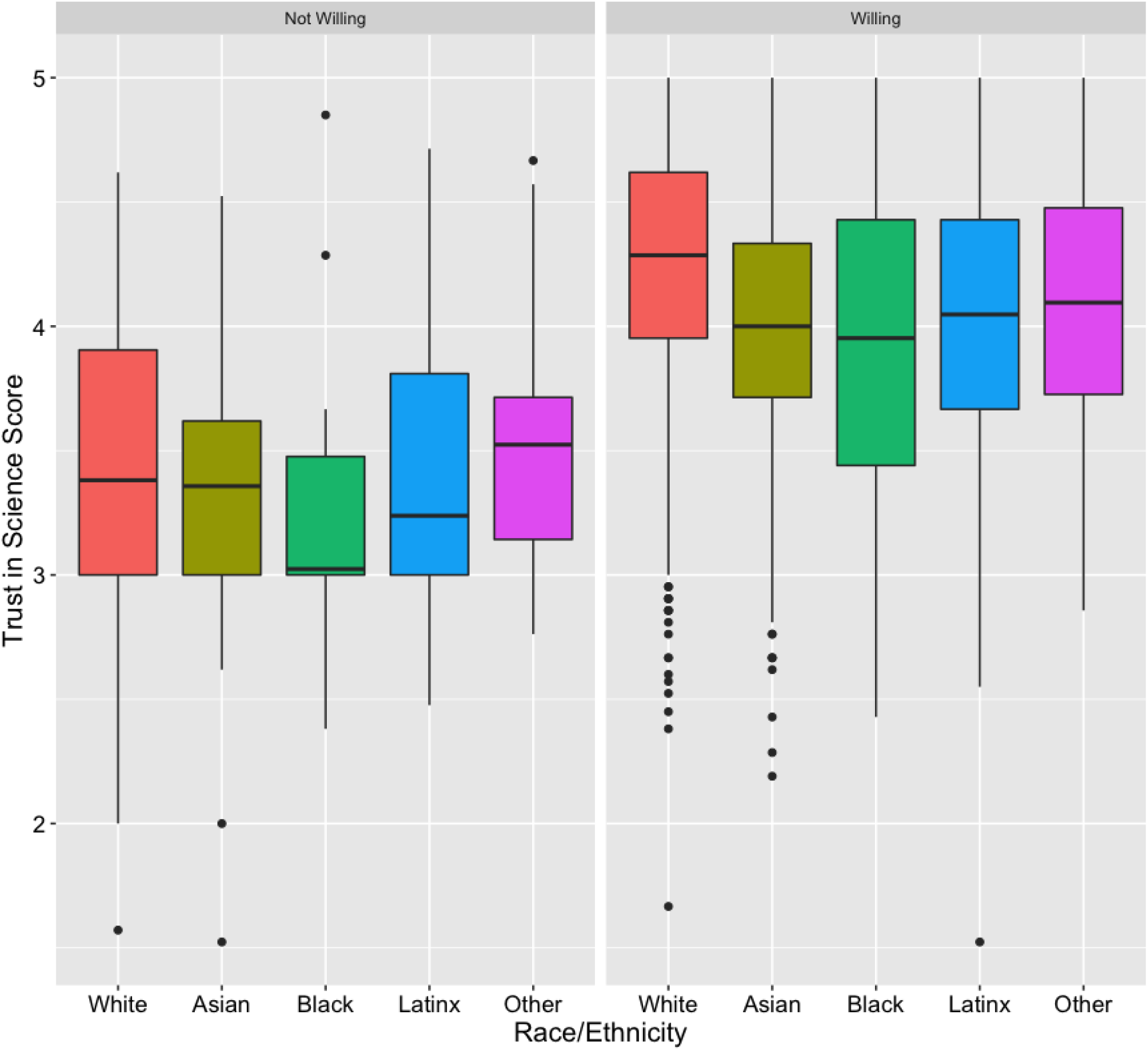
Trust in Science Scores by Racial/Ethnic Groups Stratified by Booster Willingness Status. On average, those who were willing to get a booster shot trusted science more than those who were not willing among all race/ethnicity groups.

## Discussion

While our sample is at a higher education setting, thus limiting generalizability, our results demonstrate high willingness to receive a COVID-19 booster shot. Our findings also highlight the importance of educational and motivational messages that focus on trust in science to increase willingness to get the COVID-19 booster. This is especially important in addressing previously reported COVID-19 vaccine hesitancy among African Americans.^6^ However, more research is needed to better understand the impact of cultural beliefs on booster willingness and vaccine hesitancy, especially given the historical trauma and legacy of racism African Americans have faced from the scientific/medical field. This understanding will help determine what messages and interventions to prioritize to increase booster willingness in the future.

Though much more work is needed to determine booster willingness at a national level, these findings are a first step in understanding peoples’ thoughts and opinions about COVID-19 booster shots. These results indicate the vast majority of students and staff would be willing to receive additional vaccine doses if recommended. Future research should examine the demographic correlates of actually receiving booster doses, which might be lower than willingness and intentions. Conversely, in settings where the vast majority of people are willing to get boosters, it will be important to monitor vaccination records to make sure that people do not obtain boosters more frequently than recommended, which could increase the risk of side effects without providing additional protection from COVID-19.

### Limitations

This analysis was based on a non-random sample of university students, staff, and faculty who responded to an online survey. Findings may not generalize to the general population.

## Conclusions

This study demonstrates an overall high COVID-19 booster willingness among University students, staff, and faculty and highlights the importance of educational and motivational messages that focus on trust in science to increase willingness to get the COVID-19 booster. Future research and public health efforts should be put on fostering trust among people who are wary of science and the COVID-19 pandemic.

## Data Availability

All data produced in the present study are available upon reasonable request to the authors

## Acknowledgements

This research has been funded by the University of Southern California Office of the Provost and the Keck Medicine Foundation. E. Kawaguchi is funded by the NIH T32ES013678. A. Kim is funded by the USC Environmental Health Biostatistics Core (P30 ES007048).

## References

1. Choi A, Koch M, Wu K, et al. Safety and immunogenicity of SARS-CoV-2 variant mRNA vaccine boosters in healthy adults: an interim analysis. Nat Med. 2021;27(11): 2025–2031. doi:10.1038/s41591-021-01527-y

2. Bar-On YM, Goldberg Y, Mandel M, et al. Protection of BNT162b2 Vaccine Booster against Covid-19 in Israel. N Engl J Med. 2021;385:1393–1400. doi:10.1056/NEJMoa2114255

3. Mahase E. Covid-19: Booster dose reduces infections and severe illness in over 60s, Israeli study reports. BMJ. 2021;374:n2297. doi:10.1136/bmj.n2297

4. AlShurman BA, Khan AF, Mac C, Majeed M, Butt ZA. What Demographic, Social, and Contextual Factors Influence the Intention to Use COVID-19 Vaccines: A Scoping Review. Int J Environ Res Public Health. 2021;18(17):9342. doi:10.3390/ijerph18179342

5. Cascini F, Pantovic A, Al-Ajlouni Y, Failla G, Ricciardi W. Attitudes, acceptance and hesitancy among the general population worldwide to receive the COVID-19 vaccines and their contributing factors: A systematic review. EClinicalMedicine. 2021;40:101113. doi:10.1016/j.eclinm.2021.101113

6. Khubchandani J, Macias Y. COVID-19 vaccination hesitancy in Hispanics and African-Americans: A review and recommendations for practice. Brain, Behavior, & Immunity - Health. 2021;15:100277. doi:10.1016/j.bbih.2021.100277

7. Nadelson L, Jorcyk C, Yang D, et al. I Just Don’t Trust Them: The Development and Validation of an Assessment Instrument to Measure Trust in Science and Scientists. School Science and Mathematics. 2014;114(2):76–86. doi:10.1111/ssm.12051

